# Cytokine release syndrome after treatment with immune checkpoint inhibitors: an observational cohort study of 2672 patients from Karolinska University Hospital in Sweden

**DOI:** 10.1101/2024.03.15.24304338

**Authors:** Osama Hamida, Frans Karlsson, Andreas Lundqvist, Marco Gerling, Lisa L. Liu

**Author notes:** Corresponding authors: Lisa L Liu Burström, MD, PhD, Marco Gerling, Dr. med. Contributed equally (Marco Gerling, Lisa L. Liu).

## Abstract

Immune checkpoint inhibitors (ICIs) are linked to diverse immune-related adverse events (irAEs). Rare irAEs surface first in clinical practice. Here, we systematically studied the rare irAE, cytokine-release syndrome (CRS), in a cohort of 2672 patients treated with ICIs at Karolinska University Hospital in Stockholm, Sweden. We find that the risk of ICI-induced CRS – defined as fever, negative microbiological findings and absence of other probable causes within 30 days after ICI treatment – is approximately 1%, higher than previously reported. ICI-induced CRS was often mild and ICI rechallenge was generally safe. Two out of 28 patients experienced high-grade CRS, and one was fatal. While C-reactive protein and procalcitonin were not discriminative of fatal CRS, our data suggest that the quick Sequential Organ Failure Assessment (qSOFA) score might identify high-risk patients. These data provide a framework for CRS risk assessment and motivate multicenter studies to improve early CRS diagnosis.

**Highlights:**

- Cytokine release syndrome following immune checkpoint inhibition is rare and often mild.
- Risk assessment using quick Sequential Organ Failure Assessment, but not serum CRP, can potentially detect severe cytokine release syndrome and improve treatment decisions.
- Rechallenge with immune checkpoint inhibitors after mild cytokine release syndrome is generally well tolerated.

## Introduction

Immune checkpoint inhibitors (ICI) have become a cornerstone in the treatment of a wide array of cancers [1]. Due to their increased use, immune-related adverse events (irAE) have become more prevalent. The activation of the immune system by checkpoint inhibition can lead to undesired off-target effects manifested as autoinflammatory conditions that can occur in any organ. With increasing knowledge of these potentially life-threatening side effects, clinical treatment guidelines for the common irAEs have been established [2-4]. Generally, treatment for irAEs is based on corticosteroids, while higher grades of irAE require additional immunosuppressants, and can lead to withholding or cessation of ICI treatment [3-5].

Recently, rare and potentially fatal irAEs driven by systematic hyperactivation of the immune system have become visible in clinical practice [6]. While cytokine release syndrome (CRS) is commonly associated with chimeric antigen receptor-modified T-cell (CAR-T) therapy [7, 8], emerging data supports that it may also be recognized as an irAE following treatment with ICI [6, 9-11]. Therefore, knowledge about the condition and treatment recommendations in the context of ICIs are extrapolated from CRS associated with CAR-T cell therapy. The knowledge of CRS following ICI is limited, and most studies are based on case reports and case series [6]. Mechanistically, CRS is driven by the release of interferon-gamma (IFN-γ) by activated T cells, which in turn stimulate the activation of macrophages producing proinflammatory factors, including interleukin-6 (IL-6) and tumor-necrosis factor alpha (TNF-α) [12, 13]. We have previously shown in a systematic review that CRS is rare, but potentially fatal in 10% of all cases [6]. However, factors associated with high-grade CRS have not been systematically characterized and markers allowing risk stratification for CRS patients are lacking, hindering evidence-based therapeutic decisions. Although CRS is mentioned in the latest Society for Immunotherapy of Cancer (SITC) guidelines for irAEs [4], it lacks clear diagnostic criteria and specific treatment guidelines. In this light, the management of hyperinflammatory syndromes after ICI treatment remains a clinical dilemma.

Here, we studied all patients admitted for CRS after ICI treatment at Karolinska University Hospital, Stockholm, Sweden over a ten-year period. We find that most cases of CRS are mild, and that clinical sepsis scores might outperform blood-based inflammation markers for early risk stratification. Our data provide a framework for CRS risk stratification, allowing early diagnosis of potentially fatal courses that require aggressive immunosuppressive therapy.

## Methods

We included all patients (n = 2672) with any solid cancer, treated at Karolinska University Hospital, Stockholm, Sweden from July 19^th^ 2012 until August 16^th^, 2022 who met the following criteria: 1) received at least one administration of any of the following substances: nivolumab, ipilimumab, pembrolizumab, atezolizumab, durvalumab, avelumab, cemiplimab and 2) admitted with one of the following diagnoses (ICD-codes) within 30 days from administration of the previous substances: R509 (Fever, unspecified), R508 (Other specified fever), A419 (Sepsis unspecified), Z038B (Observation/examination for suspected infectious disease), R502 (Fever caused by medical agent), A418 (other specified causes of sepsis), R651 (Sepsis according to Sepsis-3-criteria), T887X (Reaction following pharmaceutical administration, unspecified), R572 (Septic shock according to Sepsis-3-criteria). Data were acquired from the Karolinska University Hospital’s health care database, and the electronic journals of the patients matching the criteria above were manually reviewed to confirm CRS diagnosis. CRS was defined as fever (>38.0 degrees) with no positive microbiological results or other signs of viral or bacterial infection, such as positive viral panels or radiological results suggestive of any infection or any lab or radiological sign for irAE such as pneumonitis, hepatitis, and pancreatitis within 30 days from ICI administration. Eligible patients’ data were extracted manually using a pre-defined set of variables (**Tables 1, 2, and 3**). All unique patients with CRS (n = 28) were included in the study and cases of recurrent CRS after rechallenge with ICI (n = 3) were noted separately in the data set. Tumor burden was defined as having at least two metastatic lesions, a minimum tumor size ranging from 6– 10 cm (corresponding to the sum of diameters), and LDH levels ≥3 times the ULN [14].

**Table 1.**
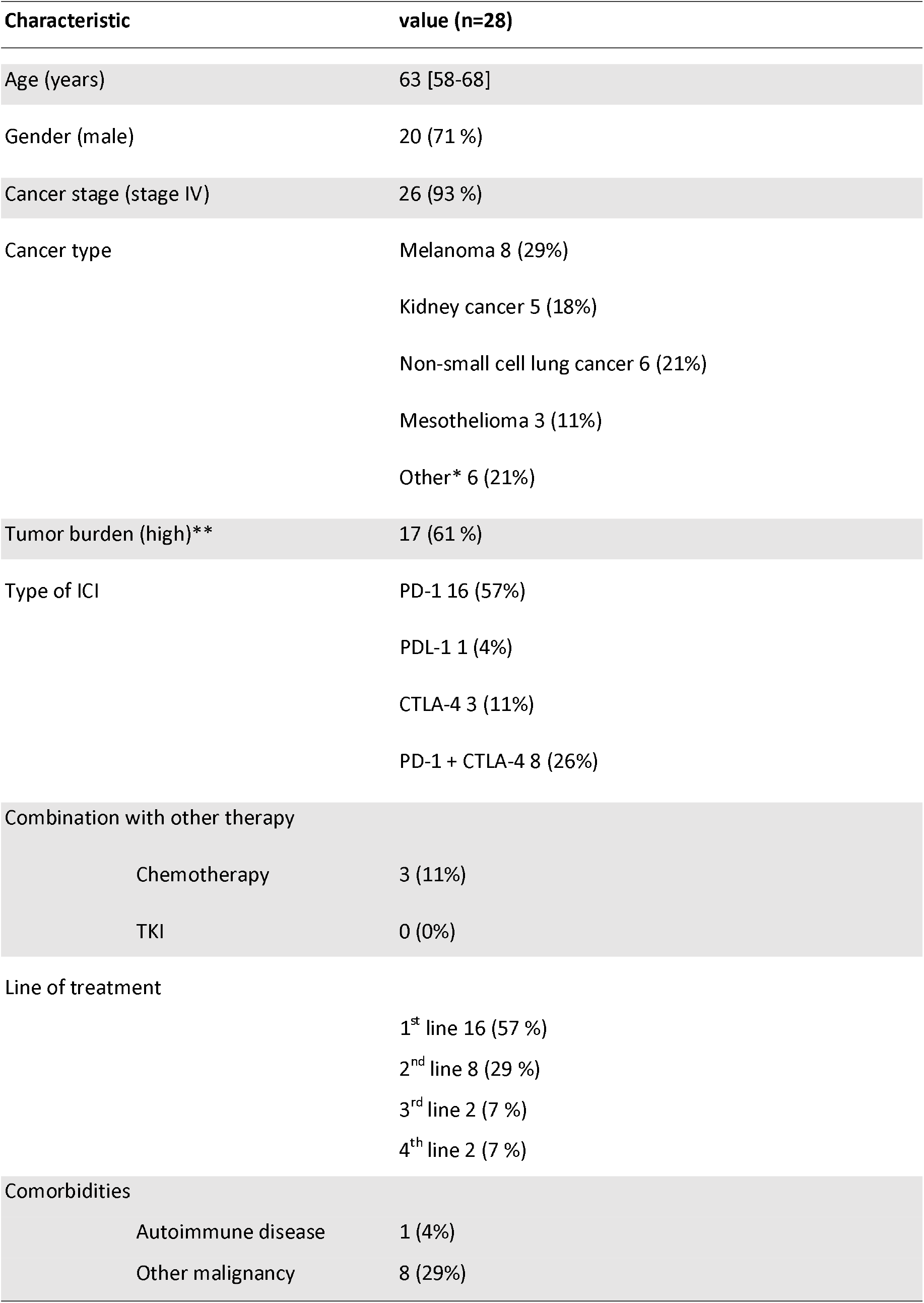
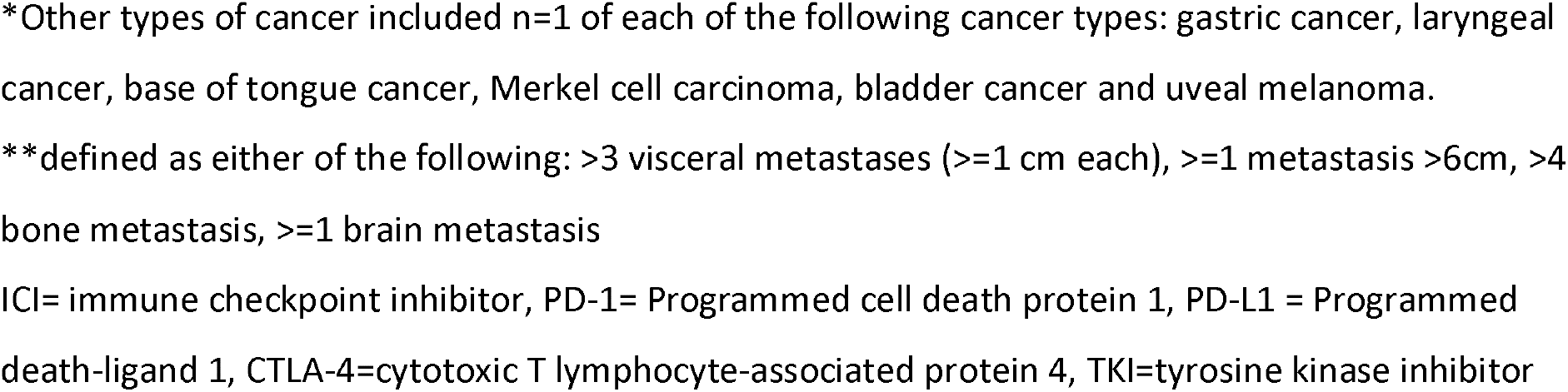
Clinical characteristics of patients with immune checkpoint-induced cytokine release syndrome.

**Table 2.**
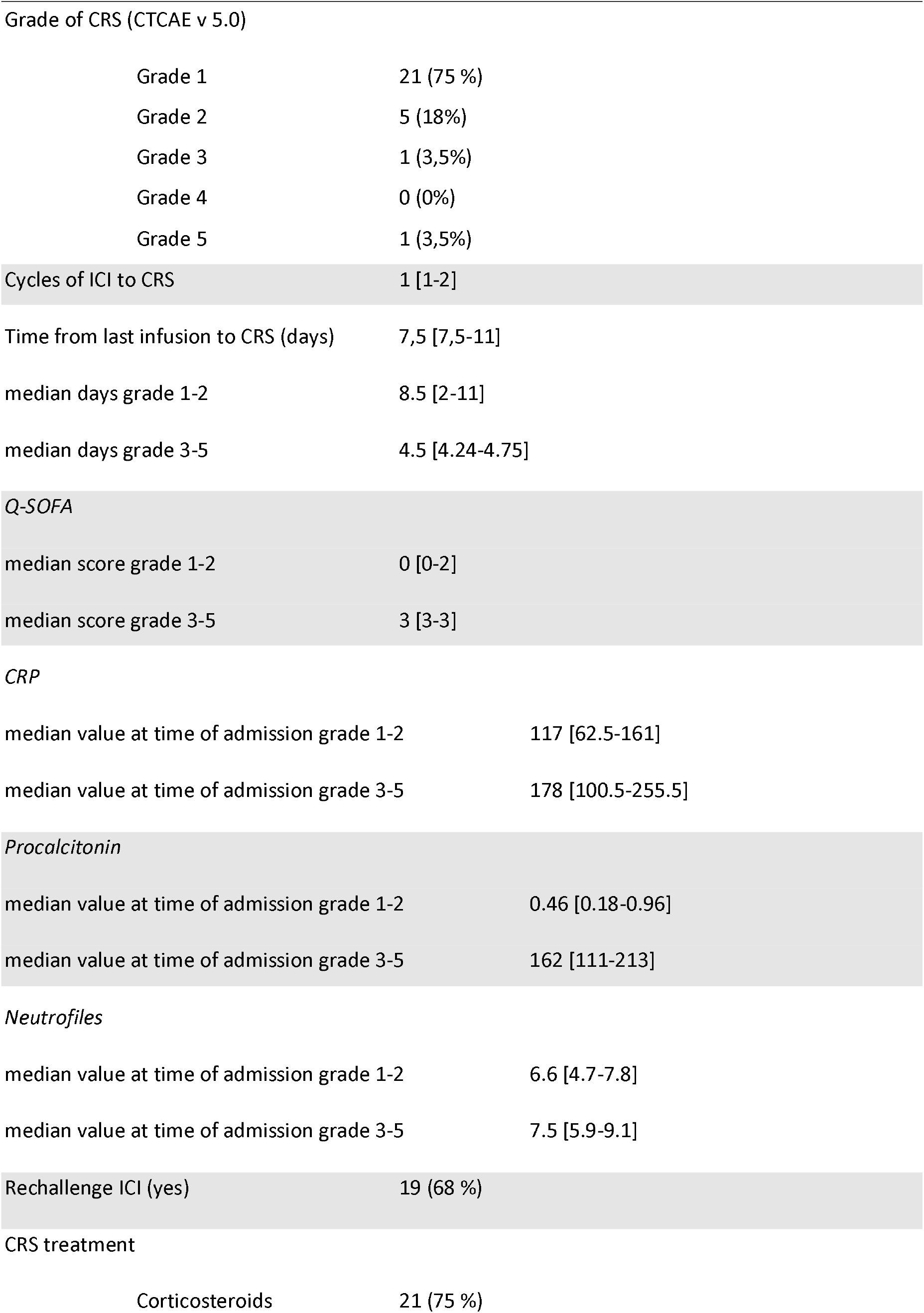
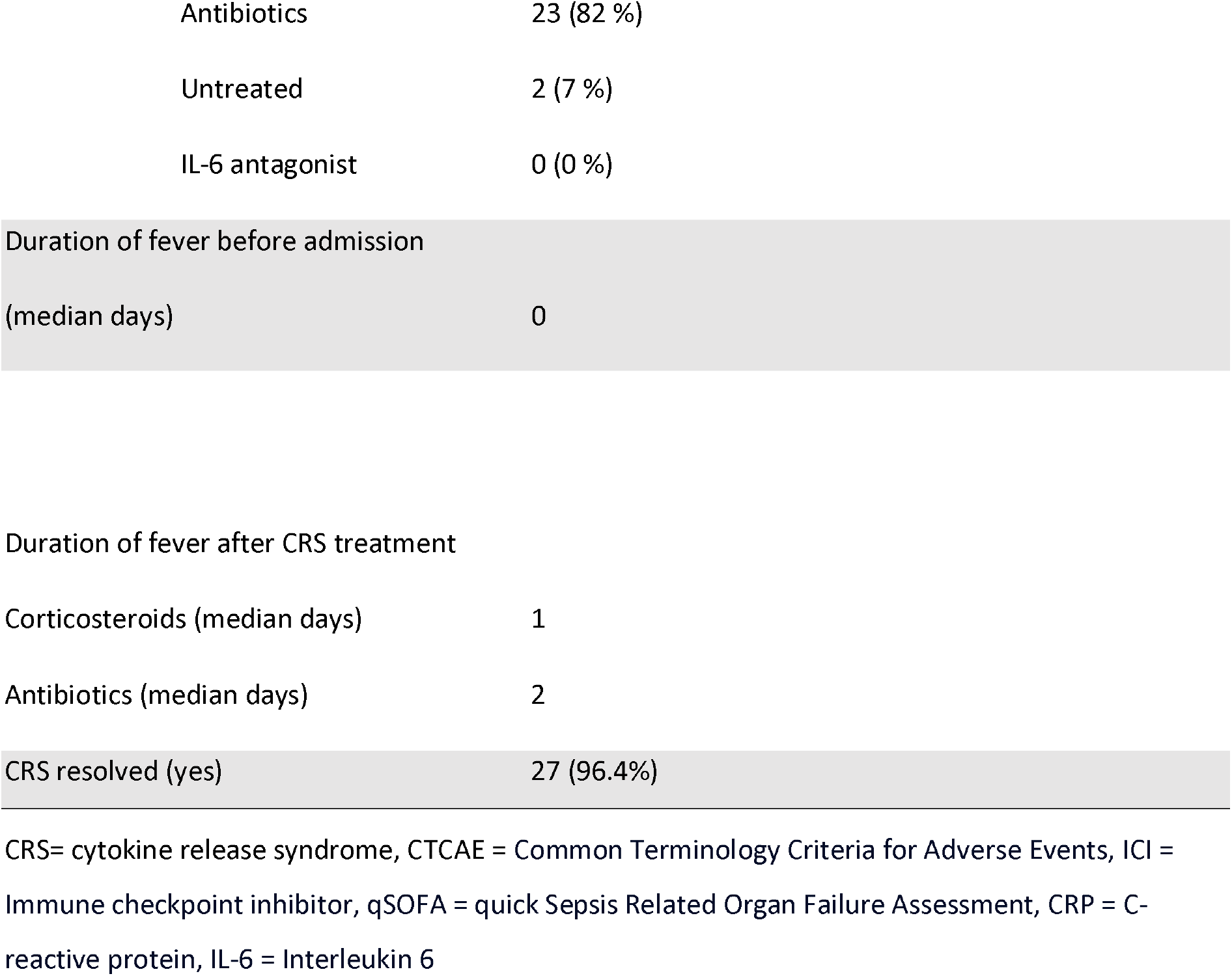
Cytokine release syndrome treatment and outcomes.

**Table 3.** Antitumoral treatment outcome following immune checkpoint inhibitor-induced cytokine release syndrome*.

|  |  |  |
| --- | --- | --- |
| 406 | CR | 1 (4,5 %) |
| 407 | PD | 15 (68 %) |
| 408 | SD | 1 (4,5 %) |
| 409 | PR | 5 (23 %) |
| 410 | Median duration of response on ICI | 12 months |

CR = Complete response, PR = Partial response, SD= Stable disease, PD = Progressive disease, ICI = immune checkpoint inhibitor
\*22 patients had evaluable radiological assessment following CRS

### Ethical considerations

The study was approved by the national Ethical Review Board (Ethical approval Dnr 2022-05600-01). Informed consent for this study was waived by the Ethical Review Board.

## Results

### Patient characteristics

In total, n = 2672 patients (n = 1445 male, n = 1227 female) received immune checkpoint blockade in the study time frame. Of these, n = 323 patients were admitted to the hospital with fever within 30 days of ICI treatment. In total, n = 28 patients (1% of the initial population of treated patients) were identified as meeting the definition of CRS within 30 days of ICI administration. Although our results align with previous reports of CRS being rare, CRS was approximately twice as common as previously estimated [6]. The median age of the patients was 59 years (Q1; Q3, 59; 69). Our cohort contained more male (n = 20) than female patients (n = 8), although we did not detect a significant difference in CRS frequency between sexes (Chi-square test, p = 0.0668). The majority of patients had stage IV disease (93 %, n = 26), and the most common cancer types were melanoma (29%), non-small cell lung cancer (NSCLC) (21%) and kidney cancer (18%). Most patients received PD-1 blockade as first-line treatment (57%, n = 16), while 26% (n=8) received dual checkpoint blockade with PD-1 + CTLA4 inhibitor (**Table 1**).

### Clinical characteristics of ICI-induced CRS

ICI-related CRS developed after a median of 7.5 days (Q1; Q3, 7.5;11) post-ICI administration and in median after the first cycle of ICI (Q1; Q3 1;2). Most patients developed grade 1-2 CRS (93 %, n = 26), whereas one patient developed grade 3 CRS and one developed grade 5 CRS (**Table 2**). Since fever is the main diagnostic criterion, it remains a clinical challenge to diagnose CRS and to dismiss other causes such as infection. Although CRP and procalcitonin were elevated upon hospital admission in our cohort, neither are specific for hyperinflammatory syndromes, and none correlated with patients developing severe CRS (**Figure 1B and C**). However, we found that a higher quick Sequential Organ Failure Assessment (qSOFA) score upon admission correlated with severe CRS (**Figure 1A**), implying qSOFA to be a simple and potentially predictive factor for severe CRS. In our cohort, both patients with grade 3 and 5 CRS had a qSOFA score of 3, while those patients developing grade 1-2 CRS scored 0-2. Due to the small sample size, it is not possible to make any statistical analysis of these data, and further studies on combined, large cohorts are warranted.

**Figure 1.**
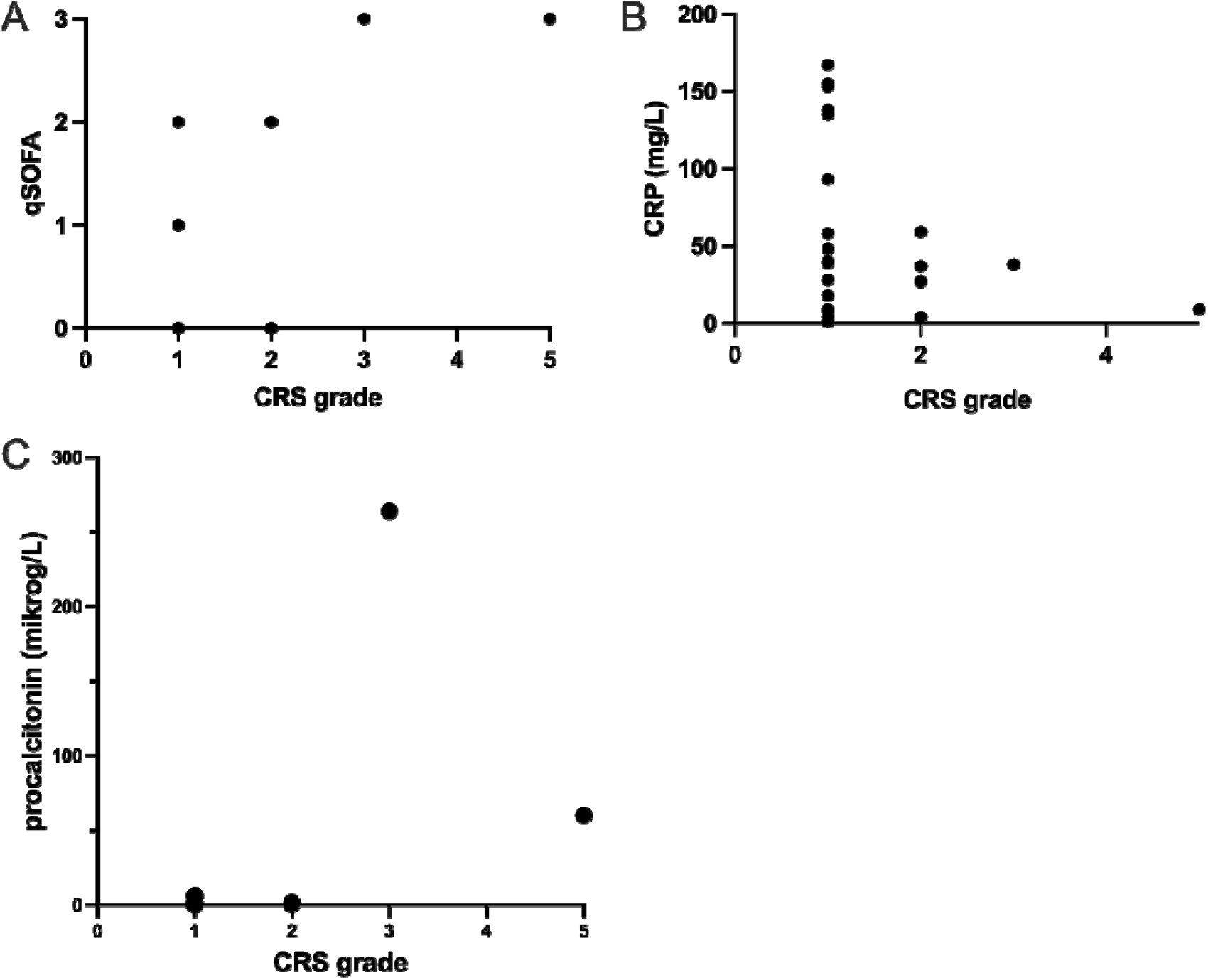
Assessment of qSOFA and laboratory tests at time of admission for cytokine release syndrome in patients receiving immune checkpoint inhibition treatment. (A) qSOFA score associated with CRS grade, n= 28. (B) Level of CRP associated with CRS grade, n = 28. (C) procalcitonin level associated with CRS grade, n=19. qSOFA: quick sepsis related organ failure assessment, CRP: C-reactive protein.

### Clinical characteristics of patients developing severe CRS

The patient developing grade 5 CRS was a male in his 80s with Merkel cell carcinoma. Five days after the first administration of avelumab, he was admitted to the hospital with a fever of39.6 degrees centigrade, CRP of 24 mg/L, leukocytes of 6,1x10^9/L and a qSOFA score of 3. No treatment with either anti-inflammatory agents or antibiotics was initiated upon admission. He had a continuous fever at 40,3 degrees centigrade on day two, CRP of 85 mg/L and started to develop hypotension. He was started on antibiotics, but his condition quickly worsened, and he passed away the day after. Due to the nature of his quickly progressing cancer and age, no escalation to the intensive care unit was made. All blood cultures taken upon admission were negative, and no other sign of infection was apparent.

The second patient developing severe CRS (grade 3), was a male in his 50s with clear cell renal cell cancer. He was started on first-line antitumoral treatment with ipilimumab/nivolumab and developed symptoms of CRS four days after the administration of the fourth cycle. He presented with a fever of 38.8 degrees centigrade and a blood pressure of 60/40 mmHg. CRP and procalcitonin levels were elevated to 284 mg/L and 264 μg/L respectively and qSOFA score 3. He was started on broad-spectrum antibiotics, i.v corticosteroids and noradrenalin upon admission. The fever resolved until the next day, to return again two days later despite ongoing corticosteroid administration and broad-spectrum antibiotics. All blood cultures taken were negative. The patient recovered and was discharged from the hospital after 21 days, whereas the fever resolved on day 11. CRP continued to be elevated after the patient was discharged and he had continuous treatment with oral antibiotics and corticosteroids.

### Management of ICI-induced CRS

The majority of patients in our cohort received corticosteroids (71%) and antibiotics (29%), while no patient received IL-6 blockade. Median time for CRS duration after administration of corticosteroids was one day.

As CRS is a potentially fatal condition, omitting ICI therapy after an episode is common. However, it is unclear whether patients developing CRS can safely be rechallenged. Previous studies have indicated that it might be possible to rechallenge patients with mild CRS [6]. In our cohort of 28 unique patients developing CRS, 68% of the patients (n = 19) were rechallenged with ICI after CRS, and only three of them (16%) developed mild CRS a second time (CRS grade 1 and 2). These data indicate that re-challenge can be considered after mild CRS.

### ICI-induced CRS and tumor responses

Whether irAEs might be associated with improved tumor responses is unclear [15-18]. However, due to the low frequency of CRS, studies on irAEs and tumor responses do not allow for the specific assessment of the prognostic impact of CRS. In our cohort, n = 22 patients had a radiological examination that allowed the assessment of tumor responses related to the ICI treatment associated with CRS. Among these, 68% (n = 15) had progressive disease, 4,5 % (n = 1) had stable disease, 23% (n = 5) had partial response and 4,5 % (n = 1) had complete remission (**Table 3**).

## Discussion

CRS is a relatively common serious adverse event in CAR-T cell therapy. However, CRS is increasingly noticed in the context of ICI treatment, but its incidence is unclear as most studies reported case series, rather than systematic analyses of large patient cohorts. Here, we report a large, single-center cohort and use pre-defined criteria to identify CRS patients.

Our data indicate that CRS occurs in approximately 1% of patients receiving ICI. Although no association between cancer type and CRS was observed, most patients had stage IV cancer, potentially suggesting an interaction between high tumor burden and CRS risk. ICI-induced CRS was most often mild, although severe and fatal cases occurred. Our study suggests that high qSOFA score is associated with severe CRS, whilst CRP, leukocyte count and pro-calcitonin might not be able to differentiate severity in the initial phase of the condition. Since patients with CRS tend to respond quickly to anti-inflammatory agents, we propose that in patients with suspected CRS and a high qSOFA score, treatment with corticosteroids should be considered and administered quickly, and potentially concomitant with antibiotics.

Our results suggest that CAR-T cell-induced CRS differs in severity from ICI-induced CRS in that ICI-induced CRS is usually mild, even in our selected population of patients requiring hospital admission. Although previous reports on ICI-induced CRS reported patient mortality of up to 10% [6], we identified one patient where CRS was the likely cause of death (3,5%). The limited number of patients in previously reported studies and diverse definitions of CRS might underly the variable frequencies in reported mortality. The potential severity of CRS urges for rapid medication with anti-inflammatory agents early in the course of the condition.

Interestingly, 70% of the patients with CRS were male, even though female patients receiving ICI were 45% of the whole cohort. This is in line with previous reports of CRS being more common in male patients [6]. Although NSCLC, renal cell cancer and melanoma are more common in men, the proportion of female versus male receiving ICI in the whole cohort were relatively equal. While autoimmune disorders are more common among women, men might be more sensitive to hyperinflammatory conditions and might be in higher risk of developing more severe disease. For instance, men have a higher risk of developing severe COVID-19 disease and higher mortality rates [19, 20]. Furthermore, one major form of estrogen, estradiol, has been shown to dampen excessive production of innate inflammatory cytokines produced by macrophages and monocytes [19]. Although no specific mechanisms are known to explain gender differences in CRS, it is evident that hormonal and genetic variabilities influence immune responses, potentially increasing susceptible to severe CRS in males.

Upon development of severe irAEs, treatment with ICI is often interrupted. In the present study, 68% of all patients with mild CRS were rechallenged and only three of them developed CRS again and none were severe, suggesting that rechallenge with ICI after mild CRS is safe. Interestingly, only one patient in our study was treated with adjuvant intent, while all others’ treatments were palliative. The patient receiving ICIs adjuvant had melanoma stage 3b and developed CRS after one cycle of nivolumab. However, a CT scan directly after the first dose of nivolumab showed metastatic disease, indicating that metastases probably were present at the time of treatment initiation. In light of this data, all patients at the Karolinska University Hospital who developed ICI-induced CRS had metastatic disease. It is therefore tempting to speculate that the presence of tumor and tumor antigens play a role in the development of CRS.

In summary, our study presents the largest single-center cohort of patients with ICI-induced CRS to date. We conclude that CRS is a rare irAE and usually mild, although fatal cases occur (3,6%). Karolinska University Hospital (KUH) is the central in- and outpatient referral center in the Stockholm Region, including emergency admissions. This means that patients with severe symptoms requiring medical assessment are referred back explicitly to KUH. Even if a patient initially chooses another hospital for presentation, transfer to KUH is usually pursued, which is captured in our cohort. Although we cannot formally measure the degree of external visits to the emergency department, this is generally uncommon, and we estimate that our data report on at least 95% of hospital visits related to complications after ICI treatment. This is important because it lets us accurately estimate the frequency of severe hyperinflammatory reactions. We posit that rechallenging with ICI after mild CRS is generally safe, although proper monitoring and management of potential CRS relapse is warranted. Potential risk factors associated with CRS are male gender and metastatic disease. The qSOFA score upon admission might aid clinical decision-making among patients developing fever after ICI administration, such that high qSOFA scores indicate potentially severe CRS, giving a clinical rationale to initiate corticosteroids promptly. As infection is difficult to rule out at the time of admission, we recommend that these patients are treated with both broad-spectrum antibiotics and anti-inflammatory agents upon admission, and if not responsive to corticosteroids, potential treatment with IL-6 inhibitors should be considered. Further studies are needed to decipher the mechanisms behind ICI-induced CRS and other potential risk factors associated with severe CRS.

## Data Availability

All data produced in the present study are available upon reasonable request to the authors; access may require a new application for ethical approval.

## List of abbreviations

CAR-T: chimeric antigen receptor modified T-cell
CRP: C-reactive protein
CRS: cytokine release syndrome
CT: computerized tomography
ICIs: immune Checkpoint Inhibitors
IFN-γ: interferon gamma
IL-6: interleukin 6 irAEs immune-related adverse events
i.v: intravenous
NSCLC: non-small cell lung cancer
OS: overall survival
qSOFA: quick Sequential Organ Failure Assessment
SITC: Society for Immunotherapy of Cancer
TNF-α: tumor necrosis factor alpha

## Author contributions

LLL and MG conceived and designed the study. OH and FK collected data. OH and LLL performed data analysis. LLL, MG, OH wrote the manuscript. AL and FK provided intellectual input. AL and FK edited the manuscript.

## Acknowledgements

The authors are grateful for support from Frida Bulukin Wilén for her help with clinical data extraction. LLL is supported by Clas Groschinsky foundation, the Swedish Cancer Society and Region Stockholm (ALF FoUI-974888). MG is supported by The Swedish Research Council, project 2018-02023.

## References

1. Bagchi, S., R. Yuan, and E.G. Engleman, Immune Checkpoint Inhibitors for the Treatment of Cancer: Clinical Impact and Mechanisms of Response and Resistance. Annu Rev Pathol, 2021. 16: p. 223–249.

2. Yin, Q., et al., Immune-related adverse events of immune checkpoint inhibitors: a review. Front Immunol, 2023. 14: p. 1167975.

3. Haanen, J., et al., Management of toxicities from immunotherapy: ESMO Clinical Practice Guideline for diagnosis, treatment and follow-up. Ann Oncol, 2022. 33(12): p. 1217–1238.

4. Naidoo, J., et al., Society for Immunotherapy of Cancer (SITC) consensus definitions for immune checkpoint inhibitor-associated immune-related adverse events (irAEs) terminology. J Immunother Cancer, 2023. 11(3).

5. Martins, F., et al., Adverse effects of immune-checkpoint inhibitors: epidemiology, management and surveillance. Nat Rev Clin Oncol, 2019. 16(9): p. 563–580.

6. Liu, L.L., et al., Systemic inflammatory syndromes as life-threatening side effects of immune checkpoint inhibitors: case report and systematic review of the literature. J Immunother Cancer, 2023. 11(3).

7. Frey, N. and D. Porter, Cytokine Release Syndrome with Chimeric Antigen Receptor T Cell Therapy. Biol Blood Marrow Transplant, 2019. 25(4): p. e123–e127.

8. Schubert, M.L., et al., Side-effect management of chimeric antigen receptor (CAR) T-cell therapy. Ann Oncol, 2021. 32(1): p. 34–48.

9. Yomota, M., et al., Cytokine Release Syndrome Induced by Immune-checkpoint Inhibitor Therapy for Non-small-cell Lung Cancer. Intern Med, 2021. 60(21): p. 3459–3462.

10. Tay, S.H., et al., Cytokine Release Syndrome in Cancer Patients Receiving Immune Checkpoint Inhibitors: A Case Series of 25 Patients and Review of the Literature. Front Immunol, 2022. 13: p. 807050.

11. Ceschi, A., et al., Immune Checkpoint Inhibitor-Related Cytokine Release Syndrome: Analysis of WHO Global Pharmacovigilance Database. Front Pharmacol, 2020. 11: p. 557.

12. Shimabukuro-Vornhagen, A., et al., Cytokine release syndrome. J Immunother Cancer, 2018. 6(1): p. 56.

13. Giavridis, T., et al., CAR T cell-induced cytokine release syndrome is mediated by macrophages and abated by IL-1 blockade. Nat Med, 2018. 24(6): p. 731–738.

14. Higuera Gómez, O., et al., “High Tumor Burden” in Metastatic Non-Small Cell Lung Cancer: Defining the Concept. Cancer Manag Res, 2021. 13: p. 4665–4670.

15. Socinski, M.A., et al., Association of Immune-Related Adverse Events With Efficacy of Atezolizumab in Patients With Non-Small Cell Lung Cancer: Pooled Analyses of the Phase 3 IMpower130, IMpower132, and IMpower150 Randomized Clinical Trials. JAMA Oncol, 2023. 9(4): p. 527–535.

16. Amoroso, V., et al., Immune-related adverse events as potential surrogates of immune checkpoint inhibitors’ efficacy: a systematic review and meta-analysis of randomized studies. ESMO Open, 2023. 8(2): p. 100787.

17. Lin, L., et al., Association between immune-related adverse events and immunotherapy efficacy in non-small-cell lung cancer: a meta-analysis. Front Pharmacol, 2023. 14: p. 1190001.

18. Bai, X., et al., Early Use of High-Dose Glucocorticoid for the Management of irAE Is Associated with Poorer Survival in Patients with Advanced Melanoma Treated with Anti-PD-1 Monotherapy. Clin Cancer Res, 2021. 27(21): p. 5993–6000.

19. Scully, E.P., et al., Considering how biological sex impacts immune responses and COVID-19 outcomes. Nat Rev Immunol, 2020. 20(7): p. 442–447.

20. Kharroubi, S.A. and M. Diab-El-Harake, Sex-differences in COVID-19 diagnosis, risk factors and disease comorbidities: A large US-based cohort study. Front Public Health, 2022. 10: p. 1029190.

